# Real-World Effectiveness of Live Attenuated vs. Inactivated Influenza Vaccines in Children

**DOI:** 10.1101/2024.12.04.24318492

**Authors:** Vera Rigamonti, Vittorio Torri, Shaun K Morris, Francesca Ieva, Carlo Giaquinto, Daniele Donà, Costanza Di Chiara, Anna Cantarutti, the CARICE study group

## Abstract

**Background and objectives:** Quadrivalent live attenuated influenza vaccines (LAIV-4) offer an alternative to inactivated influenza vaccines (IIV) for children aged 2-17 years, but data on their comparative effectiveness are limited. This study assessed vaccination rates and real-world effectiveness of LAIV-4 and IIV in preventing influenza and influenza-like illness (ILI) in Italian children during the 2022-2023 and 2023-2024 seasons.

**Methods:** We conducted a population-based cohort study of children aged 2-14 years from September 2022 to April 2024, using data from Pedianet, a pediatric primary care database of anonymized records from family pediatricians. Children vaccinated with LAIV-4 or IIV were compared to unvaccinated children. The primary outcome was any first influenza or ILI episode. Monthly vaccination incidence rates per 1,000 person-months were calculated for each vaccine type. Hazard ratios (HRs) and their 95% confidence intervals (CIs) for vaccine effectiveness (VE) were estimated using adjusted mixed-effects Cox models.

**Results:** A total of 65,545 (472,173 person-months) and 72,377 (527,348 person-months) children were included for the 2022-2023 and 2023-2024 seasons, respectively. Vaccination rates were 12.71 and 12.85 per 1,000 person-months, respectively. Compared to unvaccinated children, LAIV-4 had an overall effectiveness of 43% (95% CI, 32%-53%), while IIV effectiveness was 54% (95% CI, 46%-61%). In 2022-2023, LAIV-4 (38% [95% CI, 12%-56%]) and IIV (49% [95% CI, 37%-58%]) had comparable effectiveness. In 2023-2024, LAIV-4 (40% [95% CI, 25%-52%]) was slightly less effective than IIV (58% [95% CI, 44%-68%])(p=0.048).

**Conclusions:** An overall moderate, comparable effectiveness of LAIV-4 and IIV in preventing influenza/ILI among Italian children was observed.

**Article Summary:** A retrospective population-based cohort analysis showing moderate effectiveness of live attenuated influenza vaccines (LAIVs) in preventing influenza/influenza-like-illness in Italian children.

**What’s Known on This Subject:** There is conflicting evidence on the effectiveness of the quadrivalent live attenuated influenza vaccine LAIV (LAIV-4) in the pediatric population.

**What This Study Adds:** This population-based study assesses the effectiveness of LAIVs against influenza/influenza-like illness (ILI) among children in Italy in the post-COVID-19 influenza seasons using real-world data. Our findings document moderate protection provided by LAIVs against influenza/ILI in the 2022-2023 and 2023-2024 seasons.

**Contributors Statement Page:** Dr. Vera Rigamonti performed the statistical analysis, interpreted the results, and drafted the initial manuscript;

Dr. Vittorio Torri conceptualized and designed the artificial intelligence algorithms; Dr. Daniele Donà contributed to data interpretation;

Drs Anna Cantarutti and Costanza Di Chiara, designed the study, contributed to the analysis plan, interpreted the results, supervised the project, and contributed to the manuscript writing;

Profs. Shaun K Morris, Francesca Ieva, and Carlo Giaquinto interpreted the results and critically reviewed the manuscript for important intellectual content.

All authors reviewed, edited, and approved the final version of the manuscript, authorized its submission for publication, and agree to be accountable for all aspects of the work.

## Background

Influenza causes seasonal epidemics and occasional pandemics, leading to significant morbidity and mortality worldwide [1]. While children and adolescents generally face a lower risk of severe illness compared to infants and the elderly [2], they remain vulnerable to severe infections [3–5]. Moreover, they play a crucial role in the spread of the virus in the communities [3–8].

Vaccination is the most cost-effective strategy to prevent seasonal influenza, reducing morbidity and reducing community transmission, thus protecting vulnerable groups [9,10]. However, the prevalence of children vaccinated against influenza remains low and, in Italy, ranged from 7% to 15% during the flu seasons from 2009 to 2018 [10].

Inactivated influenza vaccines (IIV) have been the cornerstone of influenza prevention for decades. Live attenuated influenza vaccines (LAIV), administered as a nasal spray, have only recently started being extensively used as an alternative to intramuscular IIV for children aged 2-17 years. Both vaccines are currently quadrivalent and updated annually based on national and international recommendations [11].

In Italy, influenza vaccination is offered free of charge to all children aged six months to six years, although it is recommended for all pediatric age groups [12]. Two-dose influenza vaccination is recommended in children younger than nine years who haven’t received previous influenza vaccinations, regardless of the type of vaccine (LAIV or IIV); one-dose influenza vaccination is recommended for other children [12].

The immune responses elicited by these two vaccine types differ significantly, potentially impacting their effectiveness [13]. IIVs induce strong serum antibody responses, particularly IgG, but have limited capacity to induce mucosal IgA or T-cell responses. Conversely, LAIVs elicit a broader immune response, encompassing cellular, humoral, and mucosal components [14]. This broader response better mimics natural infection and may offer enhanced protection against a wider range of influenza strains, including those not closely matched to the vaccine, as well as reducing transmission [14].

Studies on the effectiveness of LAIV and IIV in children show inconsistent results [15–18]. Some comparative studies suggest superior efficacy of LAIV over IIV [3,19–21]. However, observational studies indicate mixed effectiveness of LAIVs depending on the setting and circulating strain [22–25].

This population-based cohort study aimed to describe the administration rate of LAIV-4 and IIV and assess the real-world effectiveness of LAIV-4 vs IIV in preventing influenza and influenza-like illness (ILI) episodes among Italian children and adolescents during the 2022-2023 and 2023-2024 influenza seasons when LAIV-4 became available in Italy.

## Materials and Methods

### Setting

Publicly funded primary healthcare services for children in Italy are delivered free of charge by individual Family Pediatricians (FPs) within the National Health System. This study utilized data from Pedianet (http://www.pedianet.it), a well-established electronic health record database specifically designed for pediatric care covering around 4% of the annual pediatric population [26]. Pedianet encompasses data from over 200 FPs across Italy who utilize the Junior Bit® software in their practices, providing a representative coverage of the Italian pediatric population [26].

This comprehensive database includes anonymized patient-level information, including demographics, growth parameters, diagnoses and symptoms (both ICD-9-CM coded and free text entries), medication prescriptions, and disease-based healthcare exemptions — which cover the costs of essential medical services such as diagnostics, treatments, and medications required for managing chronic conditions— and vaccination history.

In Italy, influenza vaccination is administered by FPs who participate voluntarily in the influenza vaccination program. These FPs receive reimbursement through the Special Professional Commitment Services (PPIPs) registry. Participation in the influenza vaccination program is not mandatory for FPs in Italy. We defined FPs as participants if they vaccinated at least 1.5% of their patients [10].

Data anonymization is performed in accordance with Italian regulations. Each anonymized record is assigned a unique numerical identifier. The data is then transmitted monthly to a centralized database located in Padua, Italy, for validation purposes. Inclusion in the Pedianet database is voluntary, and parents/legal guardians must provide general consent for their children’s anonymized data to be stored and used for research purposes. The Internal Scientific Committee of Società Serivizi Telematici Srl, the legal owner of Pedianet granted ethical approval of the study and access to the database.

### Study design and study cohort criteria

We conducted a retrospective observational cohort study on children aged 2 to 14 years enrolled in the Pedianet database and followed by FPs adhering to the influenza vaccination program during the observation period from September 1, 2022, to April 30, 2024. We worked on two subsequent flu seasons; the first went from September 1, 2022 to April 30, 2023, and the second from September 1, 2023 to April 30, 2024. To ensure complete data on exposure, outcomes, and covariates, we only included children who adhered to the recommended well-child visit schedule with their FP, aligning with established protocols outlined in previous research [27], and with at least one year of follow-up before entry into the cohort.

Children who recorded any influenza/ILI episode from the day of influenza vaccination (day 0) to 14 days post-vaccination were excluded.

### Exposure to influenza vaccine

Children were classified as exposed or unexposed to influenza vaccination for each season included in the study. Exposed children were further classified into those exposed to LAIV-4 and those exposed to IIV, based on whether they received a single dose of each respective vaccine type. Unexposed children were considered the reference group.

### Outcome and definitions

The primary outcome of the study was any influenza/ILI episode. Only the first seasonal influenza/ILI episode for each child was considered, as repeated influenza episodes within the same season are rare (1.6%), and influenza itself may confer immunity against future infections [28].

An influenza/ILI episode was defined based on an artificial intelligence algorithm, which used any reported clinical diagnosis of influenza (ICD-9-CM code: 487, 487.0, 487.1, 487.8), and/or free text of medical charts. A tailored Natural Language Processing algorithm to identify influenza/ILI cases from free text data was defined and validated on manually annotated data. The algorithm analyzed both the free text fields of the Pedianet database and, for patients admitted to hospitals in the Veneto region, the text of their discharge letters. These two sources were found to be complementary in identifying patients with influenza (**Supplementary materials – Description of influenza artificial intelligence algorithms**).

### Covariates

Covariates used for confounding adjustment included demographic variables (sex, age at the start of each influenza season, Italian region of birth, and deprivation index [29]), influenza vaccination status, number of influenza/ILI episodes, antibiotic therapies, and primary care visits in the epidemiological season preceding the season of interest, and comorbidities.

Children were classified as having comorbidity if they had at least one disease-based healthcare exemption for chronic complex condition (i.e., cystic fibrosis, diabetes, chronic obstructive pulmonary disease, and asthma), immunodeficiency or immunosuppressive therapy, neurological and neurocognitive conditions (including Trisomy 21), prematurity (less than 37 weeks of gestation), renal failure, congenital cardiac disease (including heart failure), or chronic liver conditions (**Supplementary materials – Table S1**).

### Statistical analysis

Administration rates of influenza vaccination stratified by vaccine type (LAIV-4 or IIV) were calculated per 1,000 person-months.

Descriptive statistics were provided, and the chi-squared test was used to assess differences between the groups of children who were unvaccinated, exposed to LAIV-4, and exposed to IIV.

Mixed-effect Cox proportional-hazards models were used to assess the effectiveness of LAIV-4 and IIV compared to unexposed children in preventing influenza/ILI, considering the FP as a random effect. Hazard ratios (HRs) and their corresponding 95% confidence intervals (CIs) were independently estimated for each vaccine type relative to the unvaccinated group. Vaccine effectiveness (VE) was then calculated as VE = (1-HR)x100.

Influenza vaccine exposure was considered a time-varying covariate to avoid immortal time bias. All models were adjusted for covariates of interest (sex, age at the start of each influenza season, Italian region of birth, deprivation index, comorbidities, and influenza vaccination status, number of influenza/ILI episodes, antibiotic therapies, and primary care visits in the epidemiological season preceding the season of interest) and for correlation within children who contributed to more than one influenza season. Follow-up began on September 1st and ended upon death, migration, transfer to an FP outside the Pedianet network, occurrence of the incident outcome, or the end of the study period, which coincided with the end of the season evaluated (April 30).

Homogeneity between the LAIV-4 and IIV effectiveness was tested using the chi-squared test [30].

Finally, sensitivity analyses were performed stratifying the main models by influenza season and replicating the main analysis on children aged 2-5 years.

All data analyses were performed using SAS statistical software, version 14.1 (SAS, College Station, TX, USA). Hypothesis tests were two-sided with a type I error of 0.05.

## Results

### Study cohort characteristics

A total of 65,545 (472,173 person-months) and 72,377 (527,348 person-months) children were included in the study for the 2022-2023 and 2023-2024 influenza seasons, respectively. The inclusion and exclusion flowchart is shown in **Figure 1**.

**Figure 1.**
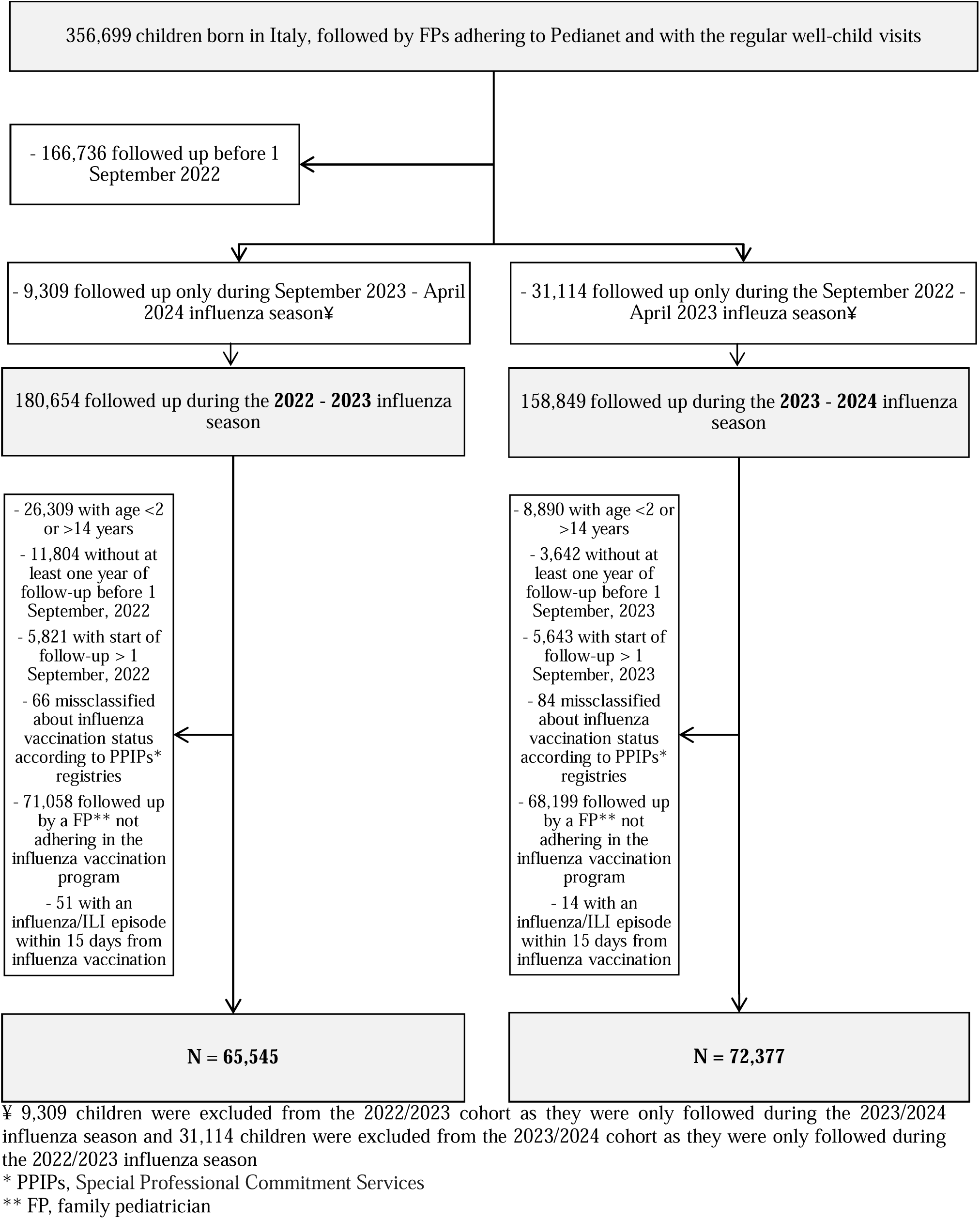
Flowchart of the study cohort. Pedianet dataset 2010-2024.

**Table 1 (Table S2 and S3)** presents the sociodemographic and clinical variables of the study population. Overall, this shows that 125,142 children were unvaccinated, 5,270 were exposed to LAIV-4, and 7,510 were exposed to IIV. Exposed children were younger, mostly born in southern Italy, and had fewer comorbidities. Comparing LAIV-4-exposed and IIV-exposed children, those in the LAIV-4 group were younger and had fewer comorbidities.

**Table 1.**
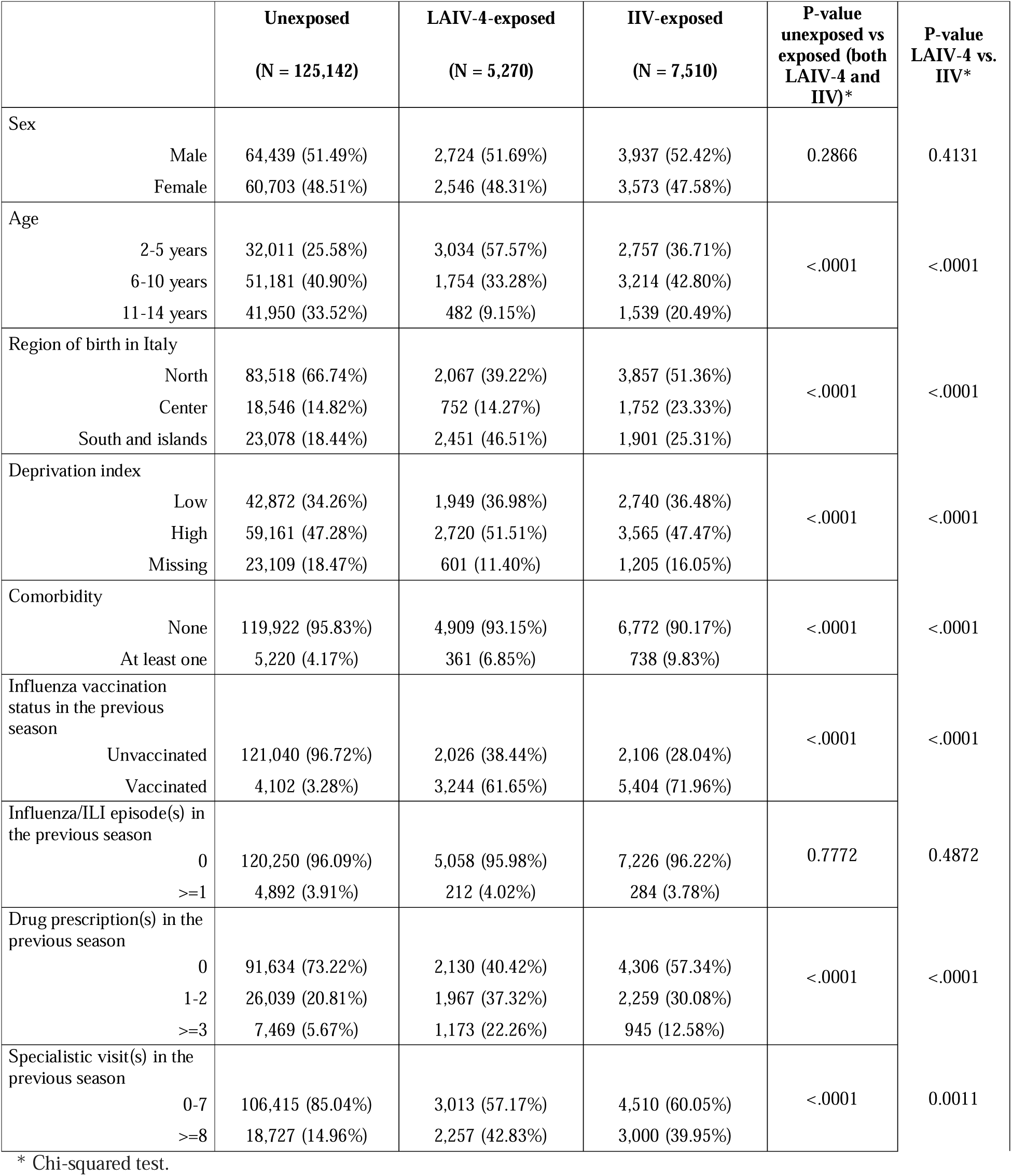
Children sociodemographic and clinical characteristics by exposure.

### Influenza vaccine administration rates by season

A total of 6,003 (12.71 per 1,000 person-months) and 6,777 (12.85 per 1,000 person-months) children were vaccinated for influenza in the 2022-2023 and 2023-2024 seasons, respectively.

In the 2022-2023 influenza season, LAIV-4 administration rate (3.56 per 1,000 person-months) was lower than that of IIV (9.15 per 1,000 person-months). Interestingly, in the 2023-2024 season, the administration rate of LAIV-4 (6.80 per 1,000 person-months) appeared to converge with that of IIV (6.05 per 1,000 person-months) (**Figure 2**).

**Figure 2.**
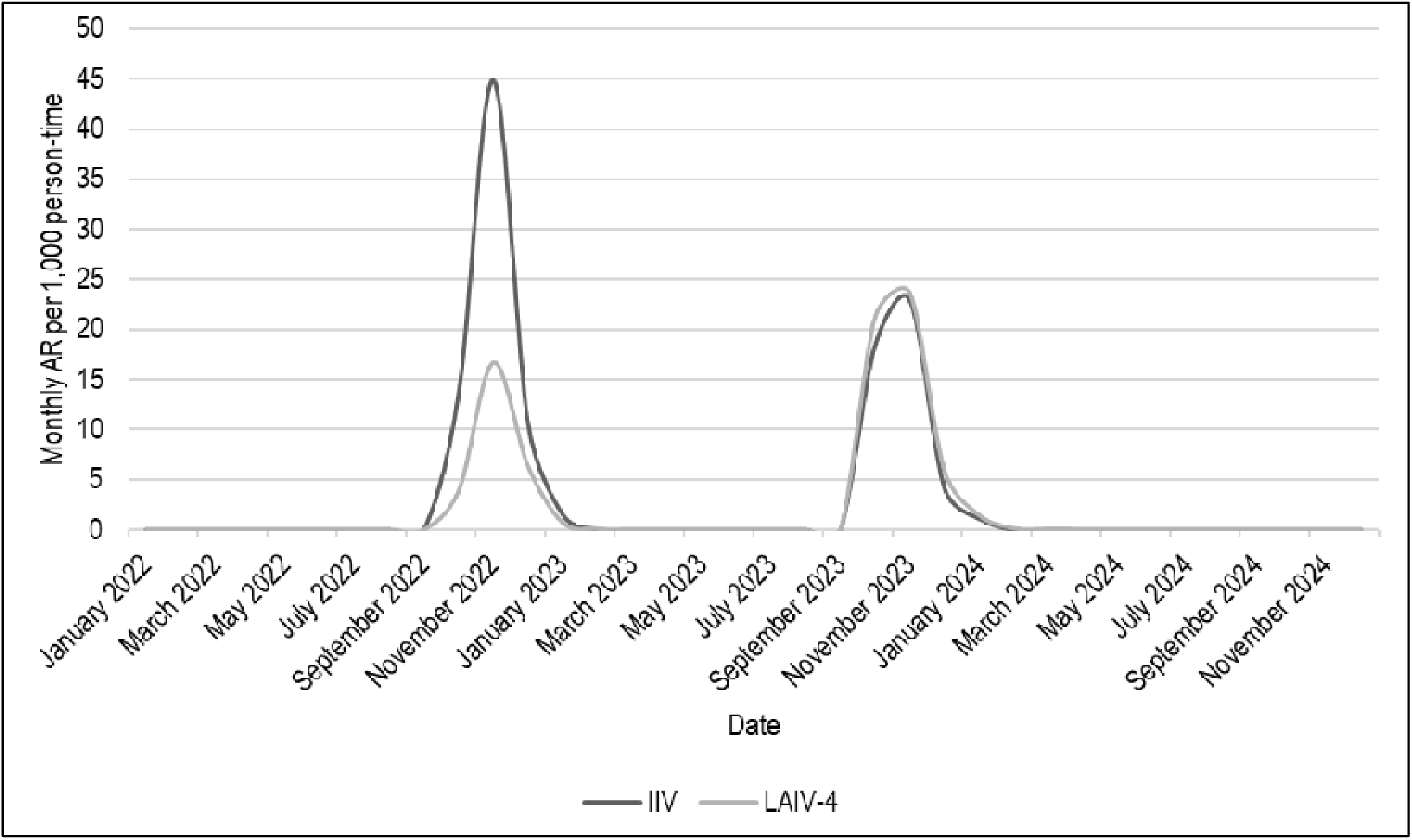
Monthly Administration Rates (AR) per 1,000 person-months of influenza vaccination among children in Italy by vaccine type.

### Effectiveness of LAIV-4 and IIV

**Figure 3** shows the effectiveness of LAIV-4 and IIV against influenza/ILI. Overall, compared to unexposed children, the effectiveness of LAIV-4 was 43% (95% CI, 32%-53%), and the effectiveness of IIV was 54% (95% CI, 46%-61%). There was no significant difference between the effectiveness of LAIV-4 and IIV in preventing influenza/ILI (p-value = 0.095).

To better understand the effectiveness of LAIV-4 and IIV against different influenza virus variants, we evaluated the effectiveness of both vaccines against influenza/ILI separately for the 2022-2023 and 2023-2024 seasons. In the 2022-2023 season, LAIV-4 (38% [95% CI, 12%-56%]) and IIV (49% [95% CI, 37%-58%]) showed comparable effectiveness (p-value = 0.374). However, in the 2023-2024 season, a slightly significantly lower effectiveness of LAIV-4 (40% [95% CI, 25%-52%]) compared to IIV (58% [95% CI, 44%-68%]) was observed (p-value = 0.048) (**Figure 3**).

**Figure 3.**
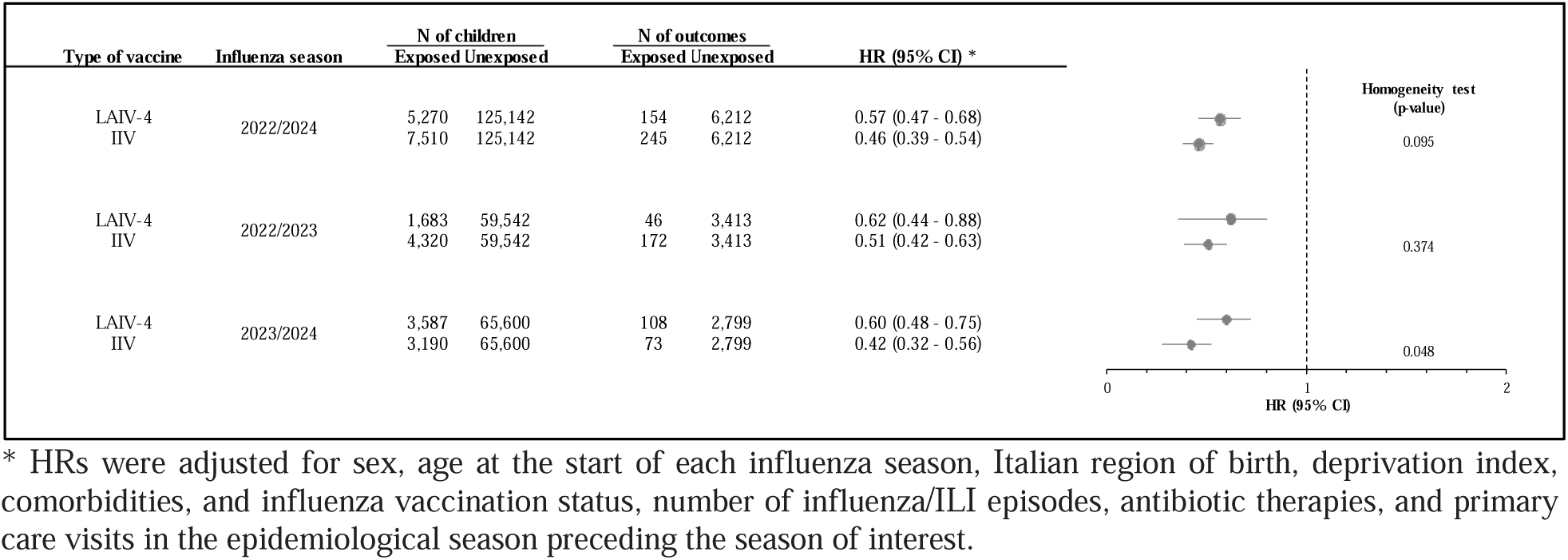
Comparison between LAIV-4 and IIV vaccine effectiveness in preventing influenza/ILI in children in Italy.

To better evaluate the impact of age on VE, we replicated the main analysis on children aged 2-5 years. The effectiveness of LAIV-4 against influenza/ILI (46% [95% CI, 32%-57%]) was comparable to that of IIV (55% [95% CI, 43%-64%]) (p-value = 0.266).

## Discussion

This nationwide study aimed to investigate the real-world effectiveness of the LAIV-4 vs IIV among children and adolescents across Italy. To the best of our knowledge, this is the first population-based study assessing the effectiveness of LAIV-4 against influenza/ILI in the Italian pediatric population using real-world data. It is also the first study in Italy to develop a Natural Language Processing algorithm for the identification of patients with influenza/ILI from free text records.

Our results strengthen and expand the evidence from other studies evaluating LAIV-4 effectiveness in the pediatric population [21,31]. We found moderate protection by LAIV-4 against influenza/ILI episodes, with VE of 38% and 40% in the 2022-2023 and 2023-2024 seasons, respectively. Previous studies evaluated the protection of LAIV-4 against influenza, observing a VE ranging from 0% to 50% in seasons 2014-2015 through 2018-2019 [17,23,24,32]. Our study found similar protection against influenza/ILI in the pediatric population compared to these results. Differences in LAIV-4 effectiveness across studies might be due to several factors, including the age and immune status of recipients and antigenic similarity between the vaccine and circulating strains [21–24,33]. Moreover, differences in influenza VE could also be attributed to varying exposure and outcome definitions among studies [31].

We observed similar effectiveness of LAIV-4 and IIV in preventing influenza/ILI in children, in accordance with findings from previous studies [17]. However, a trend emerged when analyzing by influenza season. Although VE was confirmed for both LAIV-4 and IIV in the two study seasons, during the 2023-2024 season, IIV appeared to offer slightly greater protection compared to LAIV-4. Previous analyses have yielded conflicting results due to variations in study populations and influenza strains. While some evidence showed a reduced effectiveness of LAIV compared to IIV against influenza A/H1N1pdm09 in children aged 2 to 17 years during the 2013– 2014 and 2015–2016 seasons [14], other research on healthy children aged 6 to 59 months indicated that LAIV were more effective than IIV during the 2004-2005 influenza season [34].

The study also documented an increase in LAIV-4 administration rates between the 2022-2023 and 2023-2024 seasons during the implementation of a new type of vaccine in Italy. These results predict a promising higher influenza vaccine uptake among children and adolescents, driven by vaccination campaigns based on less invasive vaccine administration. This aligns with previous research showing caregivers were more likely to accept nasal spray vaccines than injectable ones [35–36].

This study has several strengths. The use of data from Pedianet, a large population-based dataset, allowed us to assess the real-world effectiveness of LAIV-4 in Italian children. While randomized clinical trials remain the gold standard for demonstrating vaccine safety and efficacy, real-world evidence has become crucial for ongoing VE monitoring, given the influenza virus’s dynamics. Furthermore, due to the Italian-wide coverage of Pedianet, which collects all sociodemographic and clinical information, we obtained complete medical information for enrolled children, thus accounting for the impact of influenza/ILI episodes and influenza vaccination in the previous season. Moreover, the high quality of data on the influenza vaccination provided by FPs adhering to the influenza vaccination program, which reimbursed FPs for every dose of administered influenza vaccine minimizing exposure misclassification.

However, this study has several limitations. Children receiving influenza vaccination may differ from unvaccinated children in some unmeasured covariates, which results in residual confounding. The exclusion of children followed by FPs not adhering to the influenza vaccination program could also affect the study. Moreover, Pedianet only includes influenza vaccinations registered by FPs, leading to a small underreporting of vaccine coverage. Additionally, since influenza/ILI cases were defined clinically rather than confirmed by laboratory testing, outcome misclassification cannot be excluded, especially given that in Italy, the 2022–2023 and 2023–2024 influenza seasons coincided with similar surges in RSV and COVID-19. Furthermore, due to the lack of hospitalization data, we were unable to assess the effectiveness of IIV and LAIV-4 against hospitalization and severe outcomes, which are the primary goals for vaccinating children. Lastly, the 2022–2024 period was characterized by low pre-existing influenza immunity in the population following the COVID-19 pandemic. While this setting provided a unique opportunity for a more direct assessment of real-world influenza VE, it also raises questions about the generalizability of our findings to seasons with higher pre-existing immunity. However, several studies suggest that pre-existing immunity may not prevent infection but does reduce illness severity [37], supporting that the absence of pre-existing immunity should not significantly affect the generalizability of our results.

In conclusion, this population-based analysis provides real-world evidence on the moderate, comparable effectiveness of LAIV-4 and IIV against influenza/ILI among children across Italy. Further studies are needed to confirm our findings in future influenza seasons and investigate the effectiveness of LAIV-4 in preventing severe influenza/ILI in children and adolescents. Research focusing on the effectiveness of the LAIV-4 in the pediatric population is crucial for optimizing pediatric influenza vaccination campaigns to increase vaccination acceptance and uptake among children.

## Data Availability

Data produced in the present study are available upon reasonable request to the corresponding author

## Abbreviations

IIV: inactivated influenza vaccine
LAIV: live attenuated influenza vaccine quadrivalent LAIV (LAIV-4)
ILI: influenza-like illness
FP: family pediatrician
VE: vaccine effectiveness
HR: hazard ratio
CI: confidence interval
PPIP: special professional commitment service

## Acknowledgments

The authors thank all the family paediatricians collaborating in Pedianet. Eva Alfieri, Michela Alfiero Bordigato, Angelo Alongi, Biagio Amoroso, Rosaria Ancarola, Barbara Andreola, Giampaolo Anese, Roberta Angelini, Maria Grazia Apostolo, Giovanna Argo, Giovanni Avarello, Lucia Azzoni, Maria Carolina Barbazza, Patrizia Barbieri, Gabriele Belluzzi, Eleonora Benetti, Filippo Biasci, Franca Boe, Stefano Bollettini, Francesco Bonaiuto, Anna Maria Bontempelli, Matteo Bonza, Sara Bozzetto, Andrea Bruna, Ivana Brusaterra, Massimo Caccini, Laura Calì, Sonia Camposilvan, Laura Cantalupi, Luigi Cantarutti, Chiara Cardarelli, Giovanna Carli, Sylvia Carnazza, Massimo Castaldo, Stefano Castelli, Monica Cavedagni, Giuseppe Egidio Cera, Chiara Chillemi, Francesca Cichello, Giuseppe Cicione, Carla Ciscato, Mariangela Clerici Schoeller, Samuele Cocchiola, Giuseppe Collacciani, Valeria Conte, Roberta Corro’, Rosaria Costagliola, Nicola Costanzo, Sandra Cozzani, Giancarlo Cuboni, Giorgia Curia, Caterina D’alia, Vito Francesco D’Amanti, Antonio D’Avino, Roberto De Clara, Lorenzo De Giovanni, Annamaria De Marchi, Gigliola Del Ponte, Tiziana Di Giampietro, Giuseppe Di Mauro, Giuseppe Di Santo, Piero Di Saverio, Mattea Dieli, Marco Dolci, Mattia Doria, Dania El Mazloum, Maria Carmen Fadda, Pietro Falco, Mario Fama, Marco Faraci, Maria Immacolata Farina, Tania Favilli, Mariagrazia Federico, Michele Felice, Maurizio Ferraiuolo, Michele Ferretti, Mauro Gabriele Ferretti, Paolo Forcina, Patrizia Foti, Luisa Freo, Ezio Frison, Fabrizio Fusco, Giovanni Gallo, Roberto Gallo, Andrea Galvagno, Alberta Gentili, Pierfrancesco Gentilucci, Giuliana Giampaolo, Francesco Gianfredi, Isabella Giuseppin, Laura Gnesi, Costantino Gobbi, Renza Granzon, Mauro Grelloni, Mirco Grugnetti, Antonina Isca, Urania Elisabetta Lagrasta, Maria Rosaria Letta, Giuseppe Lietti, Cinzia Lista, Ricciardo Lucantonio, Francesco Luise, Enrico Marano, Francesca Marine, Lorenzo Mariniello, Gabriella Marostica, Sergio Masotti, Stefano Meneghetti, Massimo Milani, Stella Vittoria Milone, Donatella Moggia, Angela Maria Monteleone, Pierangela Mussinu, Anna Naccari, Immacolata Naso, Flavia Nicoloso, Cristina Novarini, Laura Maria Olimpi, Riccardo Ongaro, Maria Maddalena Palma, Angela Pasinato, Andrea Passarella, Pasquale Pazzola, Monica Perin, Danilo Perri, Silvana Rosa Pescosolido, Giovanni Petrazzuoli, Giuseppe Petrotto, Patrizia Picco, Ambrogina Pirola, Lorena Pisanello, Daniele Pittarello, Elena Porro, Antonino Puma, Maria Paola Puocci, Andrea Righetti, Rosaria Rizzari, Cristiano Rosafio, Paolo Rosas, Bruno Ruffato, Lucia Ruggieri, Annamaria Ruscitti, Annarita Russo, Pietro Salamone, Daniela Sambugaro, Luigi Saretta, Vittoria Sarno, Valentina Savio, Nico Maria Sciolla, Rossella Semenzato, Paolo Senesi, Carla Silvan, Giorgia Soldà, Valter Spanevello, Sabrina Spedale, Francesco Speranza, Sara Stefani, Francesco Storelli, Paolo Tambaro, Giacomo Toffol, Gabriele Tonelli, Silvia Tulone, Angelo Giuseppe Tummarello, Cristina Vallongo, Sergio Venditti, Maria Grazia Vitale, Concetta Volpe, Francesco Paolo Volpe, Aldo Vozzi, Giulia Zanon, Maria Luisa Zuccolo.

## Notes

**Conflict of Interest Disclosures (includes financial disclosures):** The authors have indicated that they have neither potential conflicts of interest nor financial relationships relevant to the article to disclose.

**Funding/Support:** This work is partially supported by CARICE (COVID-19 and Acute Respiratory Infections: the Clinical and Epidemiological changes in the pediatric population) project, which is funded by the Ministry of University and Research within the PRIN 2022 PNRR.

### Competing Interest Statement

The authors have declared no competing interest.

### Funding Statement

This work is partially supported by CARICE (COVID-19 and Acute Respiratory Infections: the Clinical and Epidemiological changes in the pediatric population) project, which is funded by the Ministry of University and Research within the PRIN 2022 PNRR.

### Author Declarations

Inclusion in the Pedianet database is voluntary, and parents/legal guardians must provide general consent for their children's anonymized data to be stored and used for research purposes. The Internal Scientific Committee of Societa' Serivizi Telematici Srl, the legal owner of Pedianet granted ethical approval of the study and access to the database.

