## Supplementary material for "Real-World Effectiveness of Live Attenuated vs. Inactivated Influenza Vaccines in Children": Supplemental-materials.docx

**Supplementary methods – Description of influenza artificial intelligence algorithms**

The algorithm for the identification of patients with influenza/ILI receives as input both the Pedianet database records, including free text fields, and, when available, discharge letters. Discharge letters are only available for children admitted to Emergency Rooms or hospitalized in the Veneto region, provided their hospitalization episode is reported in the Pedianet database.

This algorithm was originally developed using data from Pedianet between 2009 and 2020 (6’746’976 records from 283’817 unique patients). Discharge letters were available for patients hospitalized in the Veneto region from 2017 to 2020, comprising a total of 31’252 discharge letters for 15’539 patients. The subset of data related to influenza season 2017-2018 was manually annotated by clinicians and used to validate the algorithm. This subset includes 410’396 records for 117’694 unique patients, with 11’978 records positively labelled for influenza, corresponding to 10’904 unique patients. Among these, 2’646 records included a discharge letter, covering 1’364 patients, with 78 positively labelled for influenza. The remaining data were used to develop the algorithm.

To construct the algorithm, textual data were clustered, and the clusters with keywords associated with influenza were manually inspected to iteratively define the rules. In each iteration, a sample of 100 records was manually evaluated, until the number of false negatives and false positives resulted to be both lower than 5.

Below is a high-level pseudocode description of the algorithm:

**Input**:

r – pedianet record

dl – discharge letter from EHR (None if not available)

**Procedure:**

**if** icd_influenza(r.diagnosis_1.icd9) **or** icd_influenza(r.diagnosis_2.icd9) **or** icd_influenza(r.diagnosis_3.icd9):

**return** **True**

**if** r.contact_reason == ‘Vaccination’:

**return** **False**

pedianet_text = concat(r.diagnosis_1, r.diagnosis_2, r.diagnosis_3, r.discharge_diagnosis, r.admission_diagnosis, r.hosp_reason, r.contact_reason, r.symptoms_1, r.symptoms_2, r.symptoms_3)

text = concat(pedianet_text, dl.text)

sentences = split_sentences(text)

**for** s **in** sentences:

**if** identify_influenza_sentence(s):

**return** **True**

**return** **False**

In particular, the following pseudocode for the identify_influenza_sentence procedure describes the Natural Language Processing algorithm that analyzes a sentence to identify influenza/ILI:

**Input:**

s – a sentence extracted from the text

**Procedure** identify_influenza_sentence**:**

s = remove_punctuation(s)

s = lowercase(s)

s = correct_typos(s) # corrects frequent typos related to influenza/ILI

**if** is_vaccinazione(s):

**return** **False**

**if** mention_influenza(s):

**if** intestinal_influenza(s) **or** influenza_verb(s) **or** emophilus_influenza(s) **or** another_subject_influenza(s) **or** influenza_contact(s) **or** in_case_influenza(s):

**return** **False**

**if** influenza_test(s):

**if** positive_influenza_test(s):

**return True**

**else**(s):

**return False**

starting_word = find_starting_influenza_word(s)

verb = get_verb(starting_word,s)

**if** verb **is** **not** **None**:

subj = get_subject(verb,s)

**if** subj **is** **None** **or** **not** is_another_person(subj):

**return** **True**

**return** **False**

**return** **True**

**return False**

The get_verb method extracts the verb for which the mention of influenza is the object, while the get_subject method extracts the subject of the verb. These methods are implemented using the dependency parser for the Italian language provided by the Spacy Python library.

The positive_influenza_test method checks for the presence of the keyword “positivo” (positive) in sentences where an influenza test is mentioned. It also verifies that this keyword is syntactically connected to the test mention and that it is not a hypothetical result.

Most of the other methods called by the identify_influenza_sentence procedure check for specific patterns in the text. They are detailed in **Table S4**.

This set of patterns is designed to exclude cases in which there is a mention of influenza/ILI, but the patient is not actually affected by influenza/ILI.

Some examples of these false positive sentences, along with their English translations:

- Familiari con influenza stagionale (*Family members with seasonal flu*)
- Ieri ha fatto vaccino per influenza (*Yesterday he had influenza vaccine*)
- L'orario è indicativo e la tempistica in cui verra effettuata la consulenza e influenzata dalle altre urgenze in atto (*The time is approximate and the timing at which counseling will be performed is affected by other ongoing emergencies –* the verb “influenzata” (affected) in Italian is the same as “having influenza”)
- Influenzavirus A e B: negativa (*Influenzavirus A and B: negative*)

*Algorithm validation*

The algorithm was validated using gold-labelled data from the 2017-2018 influenza season.

Validation results, including true positives (TP), false positives (FP), true negatives (TN), false negatives (FN), sensitivity (TP / (TP+FN)), specificity (TN / (TN+FP)), precision (TP / (TP+FP)) and f1-score (2*precision*sensitivity / (precision + sensitivity)), are reported in **Table S5**.

The results are reported considering both the full 2017-2018 dataset and only the subset with discharge letters. For both datasets, we compared results computed considering labels derived from discharge letters, Pedianet and their combination. Considering the results on labels corresponding to the input data (i.e., discharge letters label with only discharge letters in input, Pedianet labels with only Pedianet data in input), the performance of the algorithm is excellent, both in identifying influenza from Pedianet records and in identifying it from discharge letters.

Adding the Pedianet labels to the evaluation of the algorithm with only discharge letters in input and vice-versa affects the sensitivity, demonstrating that the two sources are complementary: both allow the identification of cases of influenza that cannot be identified by the other data source.

The results with the highest number of TP are indeed those that combine Pedianet and discharge letters, and they maintain a limited number of FP and FN.

An additional validation was conducted using data from the 2022-2024 influenza seasons, to confirm the algorithm’s performance on the current study cohort. In this case, Pedianet records were manually analysed up to the identification of 100 positive and 100 negative examples. The balance between the two classes is due to the need to assess the goodness of the algorithm without manually labelling another very large dataset. The same process has been repeated for the discharge letters of the 2022-2024 season (12’069 records in total).

The results, reported in **Table S6**, are consistent with those of the 2017-2018 full dataset. Due to the limited sample size and class balance, a direct comparison between Pedianet and discharge letter labels was not performed here.

***Table S1.*** Comorbidities’ exemption codes used.

| **Disease** | **Exemption Code** |
| --- | --- |
| Cystic fibrosis | 018 |
| Diabetes mellitus | 013 |
| Chronic obstructive pulmonary disease | 024, 057 |
| Asthma | 007 |
| Congenital and acquired immunodeficiency (including HIV) and/or immunosuppressive therapy | 003, 020, 048, 050, 052 |
| Neurological and neurocognitive conditions (including Down syndrome) | 011, 017, 038, 044, 065 |
| Prematurity | 040 |
| Renal failure | 023, 061 |
| Congenital cardiac disease (including heart failure) | 002, 021 |
| Chronic liver conditions | 008, 016 |

***Table S2.*** Children sociodemographic and clinical characteristics by exposure, flu season 2022-2023.

|  | **Unexposed** | **LAIV-4-exposed** | **IIV-exposed** | **P-value*** | **P-value LAIV-4 vs. IIV*** |
| --- | --- | --- | --- | --- | --- |
|  | **(N = 59,542)** | **(N = 1,683)** | **(N = 4,320)** |  |  |
| Sex |  |  |  | 0.2038 | 0.1760 |
| Male | 30,577 (51.35%) | 854 (50.74%) | 2,276 (52.69%) |  |  |
| Female | 28,965 (48.65%) | 829 (49.26%) | 2,044 (47.31%) |  |  |
| Age |  |  |  | <.0001 | <.0001 |
| 2-5 years | 15,546 (26.11%) | 1,012 (60.13%) | 1,607 (37.20%) |  |  |
| 6-10 years | 25,153 (42.24%) | 550 (32.68%) | 1,886 (43.66%) |  |  |
| 11-14 years | 18,843 (31.65%) | 121 (7.19%) | 827 (19.14%) |  |  |
| Region of birth in Italy |  |  |  | <.0001 | <.0001 |
| North | 42,935 (72.11%) | 481 (28.58%) | 2,298 (53.19%) |  |  |
| Center | 7,333 (12.32%) | 298 (17.71%) | 847 (19.61%) |  |  |
| South and islands | 9,274 (15.58%) | 904 (53.71%) | 1,175 (27.20%) |  |  |
| Deprivation index |  |  |  | <.0001 | <.0001 |
| Low | 20,636 (34.66%) | 585 (34.76%) | 1,591 (36.83%) |  |  |
| High | 27,888 (46.84%) | 912 (54.19%) | 2,058 (47.64%) |  |  |
| Missing | 11,018 (18.50%) | 186 (11.05%) | 671 (15.53%) |  |  |
| Comorbidity |  |  |  | <.0001 | <.0001 |
| None | 57,011 (95.75%) | 1,577 (93.70%) | 3,898 (90.23%) |  |  |
| At least one | 2,531 (4.25%) | 106 (6.30%) | 422 (9.77%) |  |  |
| Influenza vaccination status in the previous season |  |  |  | <.0001 | <.0001 |
| Unvaccinated | 57,452 (96.49%) | 634 (37.67%) | 1,193 (27.62%) |  |  |
| Vaccinated | 2,090 (3.51%) | 1,049 (62.33%) | 3,127 (72.38%) |  |  |
| Influenza/ILI episode(s) in the previous season |  |  |  | 0.0527 | 0.0188 |
| 0 | 59,051 (99.18%) | 1,676 (99.58%) | 4,275 (98.96%) |  |  |
| >=1 | 491 (0.82%) | 7 (0.42%) | 45 (1.04%) |  |  |
| Drug prescription(s) in the previous season |  |  |  | <.0001 | <.0001 |
| 0 | 47,393 (79.60%) | 778 (46.23%) | 2,829 (65.49%) |  |  |
| 1-2 | 9,909 (16.64%) | 591 (35.12%) | 1,127 (26.09%) |  |  |
| >=3 | 2,240 (3.76%) | 314 (18.66%) | 364 (8.43%) |  |  |
| Specialistic visit(s) in the previous season |  |  |  | <.0001 | 0.0131 |
| 0-7 | 49,962 (83.91%) | 924 (54.90%) | 2,524 (58.43%) |  |  |
| >=8 | 9,580 (16.09%) | 759 (45.10%) | 1,796 (41.57%) |  |  |

* Chi-squared test.

***Table S3.*** Children sociodemographic and clinical characteristics by exposure, flu season 2023-2024.

|  | **Unexposed** | **LAIV-4-exposed** | **IIV-exposed** | **P-value*** | **P-value LAIV-4 vs. IIV*** |
| --- | --- | --- | --- | --- | --- |
|  | **(N = 65,600)** | **(N = 3,587)** | **(N = 3,190)** |  |  |
| Sex |  |  |  | 0.7488 | 0.9582 |
| Male | 33,862 (51.62%) | 1,870 (52.13%) | 1,661 (52.07%) |  |  |
| Female | 31,738 (48.38%) | 1,717 (47.87%) | 1,529 (47.93%) |  |  |
| Age |  |  |  | <.0001 | <.0001 |
| 2-5 years | 16,465 (25.10%) | 2,022 (56.37%) | 1,150 (36.05%) |  |  |
| 6-10 years | 26,028 (39.68%) | 1,204 (33.57%) | 1,328 (41.63%) |  |  |
| 11-14 years | 23,107 (35.22%) | 361 (10.06%) | 712 (22.32%) |  |  |
| Region of birth in Italy |  |  |  | <.0001 | <.0001 |
| North | 40,583 (61.86%) | 1,586 (44.22%) | 1,559 (48.87%) |  |  |
| Center | 11,213 (17.8092%) | 454 (12.66%) | 905 (28.37%) |  |  |
| South and islands | 13,804 (21.04%) | 1,547 (43.13%) | 726 (22.76%) |  |  |
| Deprivation index |  |  |  | <.0001 | <.0001 |
| Low | 22,236 (33.90%) | 1,364 (38.03%) | 1,149 (36.02%) |  |  |
| High | 31,273 (47.67%) | 1,808 (50.40%) | 1,507 (47.24%) |  |  |
| Missing | 12,091 (18.43%) | 415 (11.57%) | 534 (16.74%) |  |  |
| Comorbidity |  |  |  | <.0001 | <.0001 |
| None | 62,911 (95.90%) | 3,332 (92.89%) | 2,874 (90.09%) |  |  |
| At least one | 2,689 (4.10%) | 255 (7.11%) | 316 (9.91%) |  |  |
| Influenza vaccination status in the previous season |  |  |  | <.0001 | <.0001 |
| Unvaccinated | 63,588 (96.93%) | 1,392 (38.81%) | 913 (28.62%) |  |  |
| Vaccinated | 2,012 (3.07%) | 2,195 (61.19%) | 2,277 (71.38%) |  |  |
| Influenza/ILI episode(s) in the previous season |  |  |  | 0.0124 | 0.0032 |
| 0 | 61,119 (93.29%) | 3,382 (94.28%) | 2,951 (92.51%) |  |  |
| >=1 | 4,401 (6.71%) | 205 (5.72%) | 139 (7.49%) |  |  |
| Drug prescription(s) in the previous season |  |  |  | <.0001 | <.0001 |
| 0 | 44,241 (67.44%) | 1,352 (37.69%) | 1,477 (46.30%) |  |  |
| 1-2 | 16,130 (24.59%) | 1,376 (38.36%) | 1,132 (35.49%) |  |  |
| >=3 | 5,229 (7.97%) | 859 (23.95%) | 581 (18.21%) |  |  |
| Specialistic visit(s) in the previous season |  |  |  | <.0001 | 0.0007 |
| 0-7 | 56,453 (86.06%) | 2,089 (58.24%) | 1,986 (62.26%) |  |  |
| >=8 | 9,147 (13.94%) | 1,498 (41.76%) | 1,204 (37.74%) |  |  |

* Chi-squared test.

***Table S4.*** Patterns that are verified by the methods called in the identify_influenza_sentence procedure.

| **Method** | **Pattern*** |
| --- | --- |
| is_vaccinazione | vaccin(o\|azione\|azioni\|at)\|profilassi vaccinale\|vacc\b\|vaccin\b \| anti(-\|\s*)?i?nfluenz \| contro\s*(l'\s*)?influen? |
| mention_influenza | \b(sr\|s\|sdr\|sindrome)\b\s*)?(((para\|simil)(\s*-\s*\|\s*)?)?influenz(?!(at\|av)) \| \binfluenza\b \| \binfluenzavirus\b \| (?<!non\s)(sintomi\|sintomatologia) influenzal |
| intestinal_influenza | influenza intestinale |
| influenza_verb | influenzato da |
| emophilus_influenza | (emophilus\|emofilo\|influenzae\|influenziae\|haemoph)  \| h(aemophilus\|emophilus\|\.\|\s) influ |
| another_subject_influenza | epidemi(a\|e)\s*(d(i\|’) influenz\|influenzal) \| <mention_influenza> a scuola \|  <mention_influenza> in famiglia |
| influenza_contact | contatt(o\|i) con persone con <mention_influenza> |
| influenza_test | (film array per virus e batteri su tampone nasale \| \b(rna\|anf\|ricerca\|antic\|1-)\b) \| (ricerca\|tampon(e\|i)\|pcr\|sierologi(a\|e)) (virus)? influenza(le\|li)? \|  (virus)? influenza (virus?) (tipo)? (A\|B\|A e B)?(negativ\|positiv) \| (negativ\|positiv)(a\|o) (a? virus)? influenza (virus?) |
| in_case_influenza | (in caso di \| se comparsa) <mention_influenza> |
| is_another_person | madre \| padre \| mamma \| pap(a\|a\'\|à) \| genitor(e\|i) \| fratell(o\|i\|ino) \| sorell(a\|e\|ina) \| cugin(a\|o\|i\|e\|etta\|etto) \| zi(a\|o) \| nonn(i\|a\|o\|e) \| amic(a\|i\|o\|) \| parent(e\|i) \| famig?liar(e\|i) \| famiglia \| compagn(i\|a\|o)) |

* Python regular expressions notation is used in pattern definitions. Single spaces are always to be intended as \s*, even when not reported for better readability.

***Table S5.*** Validation of the algorithm for influenza identification on 2017-2018 gold-labelled data.

| **Dataset** | **Input data** | **Eval Labels** | **TP** | **FP** | **TN** | **FN** | **Sens.** | **Spec.** | **Prec.** | **F1-Score** |
| --- | --- | --- | --- | --- | --- | --- | --- | --- | --- | --- |
| 2017-2018 with discharge letters | Discharge letters | Discharge letters only | 34 | 0 | 2,612 | 0 | 100.00% | 100.00% | 100.00% | 100.00% |
| 2017-2018 with discharge letters | Discharge letters | Pedianet + Discharge letters | 34 | 0 | 2,567 | 45 | 43.04% | 100.00% | 100.00% | 60.18% |
| 2017-2018 with discharge letters | Pedianet | Pedianet | 51 | 0 | 2,595 | 0 | 100.00% | 100.00% | 100.00% | 100.00% |
| 2017-2018 with discharge letters | Pedianet | Pedianet + Discharge letters | 51 | 0 | 2,567 | 28 | 64.56% | 100.00% | 100.00% | 78.46% |
| 2017-2018 with discharge letters | Pedianet + Discharge letters | Pedianet + Discharge letters | 79 | 0 | 2,567 | 0 | 100.00% | 100.00% | 100.00% | 100.00% |
| 2017-2018 full | Pedianet | Pedianet | 11,943 | 21 | 398,425 | 7 | 99.94% | 99.99% | 99.82% | 99.88% |
| 2017-2018 full | Pedianet | Pedianet + Discharge letters | 11,943 | 21 | 398,397 | 35 | 99.71% | 99.99% | 99.82% | 99.77% |
| 2017-2018 full | Pedianet + discharge letters | Pedianet + Discharge letters | 11,971 | 21 | 398,397 | 7 | 99.94% | 99.99% | 99.82% | 99.88% |

***Table S6.*** Validation of the algorithm for influenza identification on 2022-2024 balanced sampled gold-labelled data.

| **Dataset** | **TP** | **FP** | **TN** | **FN** | **Sens.** | **Spec.** | **Prec.** | **F1-Score** |
| --- | --- | --- | --- | --- | --- | --- | --- | --- |
| 2022-2024 Pedianet sample | 100 | 0 | 100 | 0 | 100.00% | 100.00% | 100.00% | 100.00% |
| 2022-2024 Discharge letters sample | 96 | 4 | 100 | 0 | 100.00% | 96.15% | 96.00% | 97.96% |
